# Predicting Depression in Canadians with or at Risk of Diabetes: A Cross-Sectional Machine Learning Analysis

**DOI:** 10.1101/2024.02.03.24302303

**Authors:** Konrad Samsel, Mohammad Noaeen, Amrit Tiwana, Sarra Ali, Aziz Guergachi, Karim Keshavjee, Zahra Shakeri

**Author notes:** These authors contributed equally to this work and share first authorship (Konrad Samsel, Mohammad Noaeen, Amrit Tiwana).

## Abstract

Depression often goes unrecognized in individuals at risk or living with diabetes, presenting considerable challenges for primary care clinicians. Although large language models and other foundation model approaches are drawing significant attention, we systematically compared six established machine learning algorithms-Logistic Regression, Random Forest, AdaBoost, XGBoost, Naive Bayes, and Artificial Neural Networks-chosen for their reliability, interpretability, and feasibility in everyday clinical settings. By benchmarking their performance under real-world constraints, we identified key factors linked to depression risk in diabetes care, including patient sex, age, osteoarthritis, hemoglobin A1c, and body mass index. Although incomplete demographic information and potential label bias limited predictive power, our results demonstrate that a diverse set of clinical features might help pinpoint high-risk patients. They also indicate a need for longitudinal follow-up and richer clinical data to enhance model accuracy. As a practical benchmark for both clinicians and data scientists, this work suggests that machine learning–based risk stratification can improve early detection of depression and inform targeted interventions in diabetic populations.

## I. INTRODUCTION

Depression is a pervasive and often underrecognized comorbidity among individuals at risk for or living with diabetes mellitus. The bidirectional relationship between depression and diabetes not only complicates metabolic management but also significantly worsens patient outcomes-from deteriorating glycemic control to an overall decline in quality of life [1–4]. Recent studies indicate that patients with diabetes are two to three times more likely to experience depression compared to the general population [5, 6]. This concerning statistic, along with the economic and emotional burden on healthcare systems [7], calls for the need for effective screening and early intervention strategies.

In the current field of advanced artificial intelligence and generative models [8], much attention is given to increasingly complex architectures. However, clinical reality often demands methods that are interpretable, robust, and easily implementable within the constraints of routine primary care data [9–12]. While large-scale deep learning models have indeed pushed the boundaries of pattern recognition, their complexity is not always an asset in heterogeneous, real-world electronic health records (EHRs). Our work deliberately refrains from the pursuit of algorithmic novelty; instead, it focuses on using a diverse suite of established machine learning techniques as a benchmarking tool to reveal clinically actionable insights. Specifically, our study leverages routinely collected clinical data to predict depression risk among Canadians with diabetes or prediabetes. We systematically compare six supervised learning algorithms– including Logistic Regression (LR), Naive Bayes (NB), Random Forest (RF), AdaBoost (AB), XGBoost (XGB), and an Artificial Neural Network (ANN)–to identify which clinical and demographic factors best indicate depression risk. Our feature set includes patient sex, age, body mass index (BMI), hemoglobin A1c, and comorbidities such as osteoarthritis and hypertension [13, 14], all chosen for their clinical relevance.

To align technical performance with clinical applicability, we employ SHapley Additive exPlanations (SHAP) [15] to deconstruct each prediction into contributions from key features. This enables us to decompose the risk predictions into meaningful contributions from each feature, thereby translating our technical findings into insights that can inform targeted, patient-centered interventions. Importantly, while the absolute predictive metrics (e.g., AUC or F1 score) achieved by our models are modest–reflecting inherent challenges such as incomplete demographic data and potential label bias [16, 17]–they serve as a transparent benchmark for future research. Therefore, this study demonstrates that traditional and well–established models can effectively elucidate the key determinants of depression risk in diabetic populations when applied to rigorously curated EHR data. This evidence reinforces the value of established methodologies in bridging the gap between technical rigor and clinical applicability, fostering an interdisciplinary dialogue between data scientists and clinicians. The subsequent sections describe our data collection and preprocessing strategy, model development, and interpretability analyses, thereby delineating a framework for more nuanced, equitable, and actionable mental health screening in diabetes care.

## II. METHODS

### A. Data Collection and Preparation

We obtained a comprehensive dataset (*N* = 8,602) from the Canadian Primary Care Sentinel Surveillance Network (CPCSSN), a national repository of de-identified electronic medical records spanning 2003 to 2015 [10]. This cross-sectional dataset comprises 34 features capturing essential demographic attributes (e.g., age, sex), anthropometric measurements (e.g., body mass index), biomarkers (e.g., hemoglobin A1c), comorbidity diagnoses, and prescription records. To ensure clinical relevance, the cohort was restricted to patients with either a confirmed diagnosis of diabetes mellitus (type 1 or type 2) [14] or an elevated hemoglobin A1c (≥ 5.7%) indicative of prediabetes [18], yielding a final analytic sample of 6,219 individuals. Depression labels were derived from clinician-documented diagnoses, reflecting routine clinical practice.

We evaluated data quality and completeness, noting that fewer than 3% of observations had missing values. Given the minimal scope of missingness, multiple imputation by chained equations (MICE) was applied to the training portion of the data [19]. All continuous variables were normalized to mean zero and unit variance to stabilize subsequent model fitting. An 80:20 train-test split was used, and the training subset exhibited class imbalance (approximately 20% with depression). To manage this, we first removed Tomek links, where a pair (**x**_*i*_, **x**_*j*_) from opposite classes satisfies each being the other’s nearest neighbor in feature space. Removing the majority class instance in each Tomek pair eliminates borderline points that can confuse the model’s decision boundary. Next, we employed SMOTE, which synthesizes minority samples using linear interpolations: 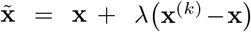, where **x**^(*k*)^ is a randomly chosen minority class neighbor of **x**, and *λ*∈ [0, 1]. This two-step balance strategy addresses the challenge in clinical data where differences between depressed and non-depressed patients can be obscured by the larger control group [20].

New variables were derived to reflect chronic disease burden, which is clinically salient for mental health outcomes. For each documented comorbidity (e.g., osteoarthritis, hypertension), we computed a duration metric by subtracting the date of diagnosis from the most recent clinical encounter, thus representing the approximate years lived with that condition. Undiagnosed cases were set to zero. Medication records were parsed into 45 binary indicators, excluding categories with fewer than ten occurrences to avoid sparse features. These feature engineering steps were motivated by evidence that longer disease duration, multiple comorbidities, and certain prescriptions may compound the risk of depression in patients with metabolic disorders.

### B. Model Development

Recognizing the inherent constraints of primary care datasets, such as modest sample sizes and heterogeneous data structures, our approach prioritizes models that strike a balance between interpretability and robust performance. While recent advances in deep learning and generative modeling have expanded the analytical toolkit, the practical limitations of routine EHR data often favor well-established, transparent methods. Accordingly, we conducted a systematic comparison of six supervised learning algorithms: LR, NB, RF, AB, XGB, and an ANN. Each algorithm was chosen based on its unique trade-off between interpretability, computational complexity, and capacity for capturing non-linear relationships. Formally, given *N* patients 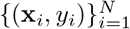, with **x**_*i*_ ∈ℝ^*d*^ representing a vector of demographics, comorbidity durations, and medication indicators, and *y*_*i*_ ∈ *{*0, 1} indicating a depression diagnosis, these methods learn a mapping *f* : ℝ^*d*^ → *{*0, 1} that generalizes to new individuals.

LR provides a straightforward linear boundary whose coefficients can serve as intuitive risk indicators, while NB uses feature-independence assumptions to yield a generative view of *P* (*y*| **x**). RF, AB, and XGB build ensembles of decision trees, enabling them to adaptively capture interactions among predictors such as age, BMI, and disease history. The ANN leverages a feedforward, multilayer structure to approximate non-linear functions, which can be helpful in capturing more complex relationships. However, as illustrated in Figure 1, larger neural networks may become over-sensitized to noise and outliers when real-world data are limited or contain label inaccuracies. Our intent was to cover a range of baseline architectures suitable for typical EHR scenarios, where advanced deep or generative models often require bigger, more uniform datasets to avoid overfitting or opaque decision boundaries. All source code for data preprocessing, model training, and interpretability analyses is publicly available^1^, facilitating replication and further refinement by both clinicians and data scientists.

**Fig. 1:**
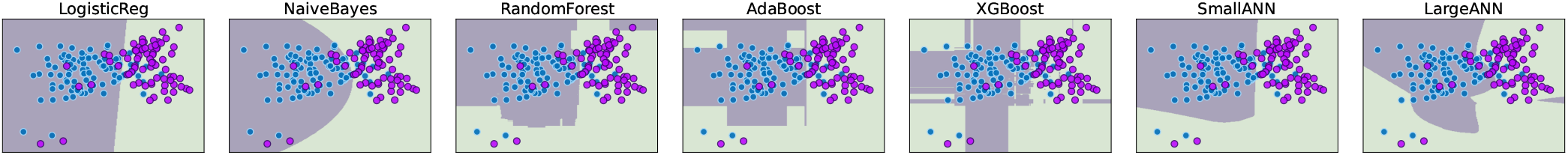
A synthetic dataset illustrating typical complexities of clinical data: correlated features, overlapping subpopulations, outliers, and label noise. Each subplot shows a learned decision boundary *f* : ℝ ^2^ →{0, 1} for a different model (Logistic Regression, Naive Bayes, Random Forest, AdaBoost, XGBoost, a smaller ANN, and a larger ANN). Simpler or ensemble methods yield stable partitions under moderate sample sizes, whereas the larger ANN overfits to minor clusters and mislabeled points. Axis labels are omitted to highlight differences in boundary shapes.

## III. Results and Discussion

Table I presents demographic and clinical attributes for 6,219 individuals who met the criteria for diabetes (62.7%) or prediabetes (37.3%). Of these, 20.7% carried a clinician-diagnosed depression, aligning with previous reports on the elevated comorbidity in metabolic conditions [5]. Notably, 50.4% were female, with hypertension emerging as the most common additional diagnosis (69.2%). To balance interpretability with completeness, we focused on hemoglobin A1c as the primary glycemic marker, given its correlation with fasting glucose and modest missingness (3%). Such choices underscore the routine data limitations in EHRs, where high correlation and partial data unavaila quently shape feature selection [17].

**TABLE I:** Patient characteristics are presented as mean [sd] ous variables or n (%) for categorical variables.

| Variable | Total (N = 6,219) |
| --- | --- |
| Age, years | 64.6 [12.4] |
| BMI, kg/m<sup>2</sup> | 30.9 [6.9] |
| Sex, female | 4,110 (52.3) |
| Hemoglobin A1c, % | 6.5 [0.9] |
| Fasting blood sugar level, mmol/L | 6.6 [1.8] |
| Total cholesterol, mmol/L | 4.6 [1.2] |
| Missing | 149 (1.9) |
| Depression (Yes) | 1620 (20.6) |
| Hypertension (Yes) | 5,439 (69.2) |
| Years living with hypertension | 2.1 [2.5] |
| Osteoarthritis (Yes) | 2,628 (33.4) |
| Years living with osteoarthritis | 2.1 [2.5] |
| COPD (Yes) | 828 (10.5) |
| Years living with COPD | 1.8 [2.0] |
| Diabetes (Yes) | 5,139 (65.4) |
| Years living with diabetes | 2.8 [2.5] |
| Take at least one hypertension medication | 6,042 (76.9) |
| Take at least one corticosteroid medication | 2,298 (29.2) |

### A. Model Benchmarking and Interpretability

Among the six models evaluated, XGBoost emerged as the top performer, achieving an AUC of 0.64 and a weighted average F1 score of 0.72 on the held-out test set. Other methods demonstrated AUCs ranging from 0.54 to 0.64. Although these performance metrics may appear modest relative to more complex deep learning architectures, they are in line with similar studies using routine EHR data for depression screening, where challenges such as imprecise labeling and the absence of detailed psychosocial measures are common [17]. The high precision in identifying non-depressed cases, alongside lower recall for the depressed minority, reflects the intrinsic class imbalance (20.6% depression rate) and the fact that some depressive conditions may remain undocumented or embedded within unstructured clinical notes. Despite these constraints, the consistent detection of several core risk factors, such as BMI, hemoglobin A1c, osteoarthritis, and younger age, suggests that even traditional machine learning algorithms can highlight clinically meaningful patterns in cross-sectional EHR data. This is important since purely deep or generative models may not always yield decisive gains when exposed to the sparse, heterogeneous records found in primary care [21]. Moreover, these reproducible but modest findings provide a transparent benchmark for researchers seeking to refine depression detection methods, underscoring two key points:

#### Label Reliability and Missing Psychosocial Variables

Depression is frequently underdiagnosed or documented in free-text notes rather than structured fields, suggesting that the actual prevalence may exceed the nominal 20.6%. The absence of standardized patient-reported outcomes or validated screening instruments (e.g., PHQ-9) further limits the model’s sensitivity and specificity [22]. These factors highlight the need for enhanced data collection protocols to improve the reliability of automated depression detection.

#### Cross-Sectional Data and Disease Trajectory

The cross-sectional design of our study precludes tracking temporal changes in glycemic control, comorbidities, or mental health status. Prior research indicates that fluctuations in metabolic parameters may precipitate or exacerbate depressive symptoms [23]. Incorporating longitudinal data could provide a more accurate representation of depression onset and progression, potentially improving predictive performance beyond the intermediate AUC values (around 0.64) observed in our study [21].

### B. Feature Importance and Clinical Implications of XGBoost

Figure 2 highlights two essential aspects of the XGBoost model’s behavior on the test set. The upper panel confusion matrix (1,244 test instances) shows 795 true negatives (non-depressed) and 96 true positives (depressed), with relatively few false positives (198) corresponding to a precision of 0.33. However, a recall of 0.38 indicates that 150 depressed patients were missed (false negatives). From a clinical standpoint, this balance may be acceptable as an initial risk stratification tool, given the well-documented challenges of accurately capturing mental health conditions in EHRs.

**Fig. 2:**
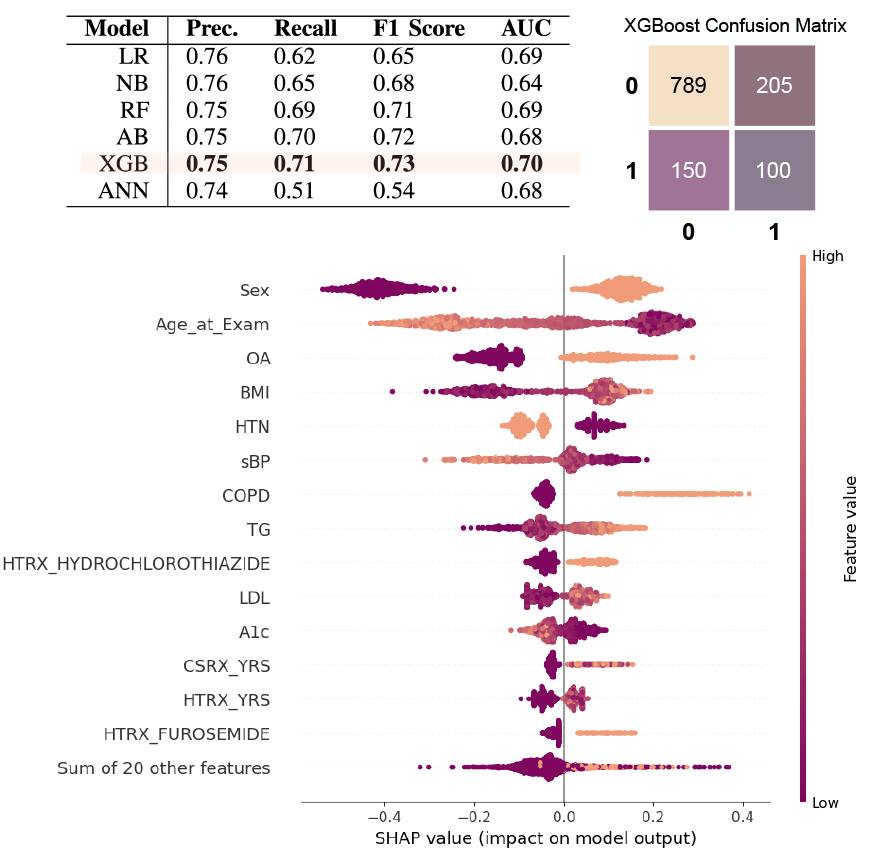
Performance metrics for six classifiers (left), XGBoost confusion matrix (top right), and SHAP summary plot (bottom). The matrix shows correctly (TN, TP) and incorrectly (FN, FP) classified patients. In the SHAP plot, each dot’s horizontal position indicates how much that feature pushes the model toward (positive) or away from (negative) a depression prediction, with purple denoting lower feature values and orange denoting higher values.

The lower panel is a SHAP summary plot, which explains feature-level contributions to the model’s predicted log-odds of depression. Specifically, for each instance **x**_*i*_ and feature *j*, the SHAP value *ϕ*_*j*_(**x**_*i*_) quantifies how much *x*_*ij*_ shifts the model’s output relative to a baseline: logi 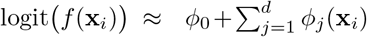, where *ϕ*_0_ is the average prediction on the training set. Positive SHAP values push the prediction toward ‘depressed’, negative values do the opposite, and the color scale (purple to orange) reflects each patient’s actual feature value. ‘Sex’ (female) and ‘Age at Exam’ (younger) emerged as influential, suggesting that female sex and earlier onset of metabolic conditions may elevate psychosocial burdens [24, 25]. Osteoarthritis (‘OA’) and BMI also significantly increased depression risk, likely reflecting pain, reduced mobility, and social constraints. Systolic blood pressure (‘sBP’) and antihypertensives (e.g., ‘`HTRX HYDROCHLOROTHIAZIDE`’) further emphasize the link between cardiovascular strain and mood disturbances. Although hemoglobin A1c did not dominate on its own, it contributed synergistically with factors like high BMI, reinforcing that depression risk in diabetic and prediabetic populations arises from multifaceted metabolic profiles rather than glycemic control alone.

Despite these clinically coherent findings, the 155 false negatives emphasize a need for improved data capture, through standardized mental health tools or more frequent longitudinal assessments, to enhance recall. Nonetheless, the SHAP interpretation demonstrates how a well-calibrated machine learning model can provide nuanced, patient-level insights into depression risk, thereby informing targeted interventions in real-world primary care settings.

### C. Limitations

Although this study leveraged real-world CPCSSN data to demonstrate the feasibility of depression risk modeling in a diverse adult population [10], several limitations remain. The cross-sectional design precludes capturing temporal changes or identifying whether evolving glycemic or comorbidity profiles trigger depressive symptoms. Reliance on clinician-documented diagnoses introduces potential label bias, as subclinical or undiagnosed cases may be overlooked; integrating standardized tools (e.g., PHQ-9) could improve label reliability. Finally, sparse demographic data restricted sub-group analysis, limiting insights into racial or socioeconomic disparities. Despite these constraints, the findings provide a valuable benchmark for future efforts to refine and expand depression detection in patients with or at risk of diabetes.

## IV. CONCLUSION

This study established a foundational benchmark for depression detection in individuals with or at risk of diabetes using routinely collected electronic health records. While XGBoost demonstrated the strongest performance (AUC ≈ 0.64), our SHAP analyses revealed that non-engineered attributes such as sex, age, osteoarthritis status, A1c, and BMI drove much of the predictive signal. These findings highlight that even standard EHR fields can flag mental health vulnerabilities, yet they also reflect the limited scope of cross-sectional data and potential label bias. Integrating longitudinal observations, richer demographic variables, and validated screening tools would likely enhance both sensitivity and specificity. As machine learning gains traction in clinical workflows, balancing model interpretability, robust data collection, and domain-guided feature selection remains paramount to realizing effective, equitable mental health support for patients facing metabolic risks.

## Data Availability

All data used are available online at https://cpcssn.ca/ website upon reasonable request.

## Footnotes

1 https://github.com/tiwanaam/mlforhealthdata

